# Epigenetic drift association with cancer risk and survival, and modification by sex

**DOI:** 10.1101/2021.03.16.21253762

**Authors:** Chenglong Yu, Ee Ming Wong, Jihoon Eric Joo, Allison M. Hodge, Enes Makalic, Daniel Schmidt, Daniel D. Buchanan, Gianluca Severi, John L. Hopper, Dallas R. English, Graham G. Giles, Melissa C. Southey, Pierre-Antoine Dugué

## Abstract

To investigate age- and sex-specific DNA methylation alterations related to cancer risk and survival, we used matched case-control studies of colorectal (*N*=835), gastric (*N*=170), kidney (*N*=143), lung (*N*=332), prostate (*N*=869) and urothelial (*N*=428) cancers, and mature B-cell lymphoma (*N*=438). Linear mixed-effects models were conducted to identify age-, sex- and age-by-sex-associated methylation markers using a discovery (controls) - replication (cases) strategy. Replication was further examined using summary statistics from Generation Scotland (GS). Associations between replicated markers and risk of and survival from cancer were assessed using conditional logistic regression and Cox models (hazard ratios [HR]), respectively. We found 32,659, 23,141 and 48 CpGs with replicated associations for age, sex and age-by-sex, respectively. The replication rates (GS summary data) for these CpGs were 94%, 86% and 91%, respectively. Significant signals for cancer risk and survival were identified at some individual age-related CpGs. There was a strong negative trend in the association between epigenetic drift and risk of colorectal cancer. Two CpGs overlapping *TMEM49* and *ARX* genes were associated with survival of overall (HR=0.91, P=7.7×10^−4^) and colorectal (HR=1.52, P=1.8×10^−4^) cancer, respectively, with significant age-by-sex interaction. Our results may provide markers for cancer early detection and prognosis prediction.

**Simple Summary:** Ageing is the strongest cancer risk factor, and men and women exhibit disparate risk profiles in terms of incidence and survival. DNA methylation is known to strongly vary by age and sex. Epigenetic drift refers to age-related DNA methylation changes and the tendency for increasing discordance between epigenomes over time, but it remains unknown to what extent the epigenetic drift might contribute to cancer risk and survival. The aims of this study were to identify age-associated, sex-associated and sexually dimorphic age-associated (age-by-sex-associated) DNA methylation markers and investigate whether age- and age-by-sex-associated markers are associated with cancer risk and survival. Our study, which used a total of 3,215 matched case-control pairs with DNA methylation in pre-diagnostic blood, is the first large study to examine the association between sex-specific epigenetic drift and cancer development and progression. The results may be useful for cancer early diagnosis and prediction of prognosis.

## 1. Introduction

Ageing is the strongest risk factor for cancer overall, and for the majority of individual cancer types [1]. Advanced age is typically associated with increased cancer risk and reduced cancer survival [2-4]. Males and females exhibit disparate risk profiles in terms of incidence and survival [5-10], e.g. most cancers with a clear sex difference affect men more than women, with incidence rates ranging from 1.3:1 for Hodgkin lymphoma to 4.9:1 for oropharynx and tonsil cancer [10]. Authors of several studies [11-15] have investigated age- and sex-specific incidence and mortality trends using population-based data. However, the mechanisms underlying the observed age-by-sex interplay are not fully understood. DNA methylation patterns are known to strongly vary by age [16, 17] and sex [18, 19]. Therefore, a key question of interest is whether DNA methylation alterations, and in which genes, show sex-specific differences during the ageing process and whether these are associated with development and progression of cancer.

Epigenetic drift is the tendency for increasing discordance between epigenomes over the lifetime [20]. Larger and higher genome-coverage studies have confirmed that the DNA methylation landscape of normal cells changes substantially with age [21], but it remains unknown to what extent the epigenetic drift might contribute to cancer risk and survival. Many authors, including Horvath and colleagues [22-24], have investigated correlations between DNA methylation and ageing, and found that males have increased DNA methylation-based age acceleration (i.e. a ‘faster ticking’ epigenetic clock) relative to females [25]. This finding was consistently reproduced in other studies [26, 27]. We and others previously found that these epigenetic aging measures were associated with risk of and survival from specific types of cancers, but the contribution of the epigenetic drift as a whole has received less attention [28-32]. A recent study [33] has identified genome-wide methylation sites at which there was chronological age-by-sex interaction and found that most of them were on the X chromosome. To the best of our knowledge, however, no study has specifically investigated whether sex-specific DNA methylation alterations related to aging are associated with cancer risk and survival.

In this study, our aims were i) to identify age-associated and sex-associated DNA methylation markers; ii) to identify sexually dimorphic age-associated (age-by-sex-associated) DNA methylation markers; iii) to investigate whether age- and age-by-sex-associated methylation markers are associated with cancer risk and survival.

## 2. Results

### 2.1. Characteristics of study samples

The characteristics of the Melbourne Collaborative Cohort Study (MCCS) samples used in this study are presented in Table 1. As shown in Table 1, controls were matched to cases on age at blood draw, sex, country of birth (Australia/New-Zealand, Greece, Italy, and UK/other) and sample type (peripheral blood mononuclear cells, dried blood spots and buffy coats). Compared with controls, cases were more frequently past and current smokers, and had greater smoking pack-years. The characteristics of the samples for each of the seven individual cancer case-control studies (colorectal, gastric, kidney, lung, prostate and urothelial cancers, and mature B-cell lymphoma) nested within the MCCS are described in Supplementary Table 1. For the lung cancer study, controls were also matched on smoking history at the time of blood collection.

**Table 1.** Characteristics of the participants in the MCCS sample included in the analysis.

| Characteristics | MCCS sample: *N* = 3,215 matched pairs |  |
| --- | --- | --- |
|  | Cases of overall cancer | Controls |
| Age at blood draw, median [IQR] | 61.5<br>[54.9-66.1] | 61.4<br>[54.5-65.9] |
| Sex: |  |  |
| male, *N* (%) | 2,237 (69.6%) | 2,237 (69.6%) |
| female, *N* (%) | 978 (30.4%) | 978 (30.4%) |
| Country of birth: |  |  |
| Australia/New-Zealand, *N* (%) | 2,162 (67.2%) | 2,148 (66.8%) |
| Greece, *N* (%) | 334 (10.4%) | 347 (10.8%) |
| Italy, *N* (%) | 505 (15.7%) | 503 (15.6%) |
| UK/other, *N* (%) | 214 (6.7%) | 217 (6.7%) |
| Blood sample type: |  |  |
| dried blood spots, *N* (%) | 2,198 (68.4%) | 2,198 (68.4%) |
| peripheral blood mononuclear cells, *N* (%) | 817 (25.4%) | 817 (25.4%) |
| buffy coats, *N* (%) | 200 (6.2%) | 200 (6.2%) |
| Smoking status: |  |  |
| current, *N* (%) | 490 (15.2%) | 468 (14.6%) |
| former, *N* (%) | 1,349 (42%) | 1,285 (40%) |
| never, *N* (%) | 1,376 (42.8%) | 1,462 (45.5%) |
| Smoking pack-years, median [IQR] | 5<br>[0-31.4] | 2.5<br>[0-28] |
| Body mass index (kg/m<sup>2</sup>), median [IQR] | 26.9<br>[24.5-29.6] | 26.9<br>[24.5-29.4] |
| Alcohol consumption (g/day), median [IQR] | 5.6<br>[0-19.7] | 4.3<br>[0-19] |

### 2.2. Identification of age-, sex- and age-by-sex-associated methylation CpGs

The discovery phase used participants selected as controls in the methylation case-control studies, and identified 48,295 age-associated and 26,331 sex-associated (P<1.0×10^−7^) methylation sites in Models 1.1 and 1.2 (without and with adjustment for smoking, alcohol consumption and body mass index [BMI], respectively; for methodology details, see Materials and Methods, and Figure 1). We also detected 73 methylation sites at which there was a significant interaction (P<1.0×10^−7^) between age and sex using Models 2.1 and 2.2 that included an age×sex interaction term. Among these markers, we replicated, using participants who were selected as cases: 32,659 age-associated CpGs (32,371 autosomal and 288 X-linked) (P<1.0×10^−6^, 68%) (Supplementary Table 2), 23,141 sex-associated CpGs (12,791 autosomal and 10,350 X-linked sites) (P<1.9×10^−6^, 88%) (Supplementary Table 3), and 48 age-by-sex-associated CpGs (4 autosomal and 44 X-linked sites) (P<6.9×10^−4^, 66%) (Supplementary Table 4). Regression coefficients of replicated associations for age, sex and age-by-sex in the discovery and replication sets are presented in Figure 2. Associations of methylation M-values for these 48 CpGs showing age-by-sex interaction are shown in Supplementary Figure 1 (discovery set).

**Figure 1.**
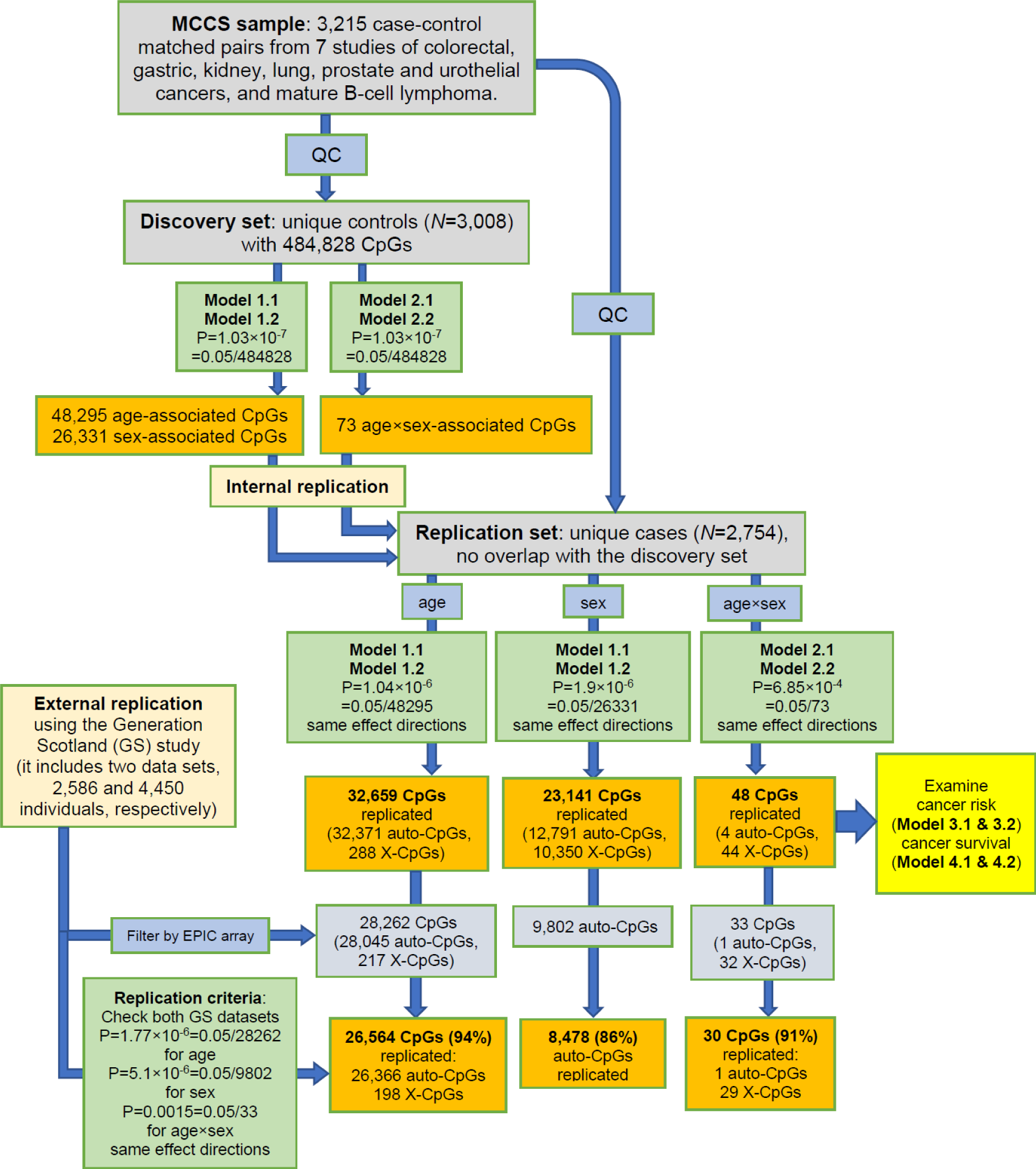
A flowchart of the discovery and replication strategy for the study. GS indicates Generation Scotland study. Auto-CpG and X-CpG indicate autosomal and X-linked CpG, respectively. The GS study reported sex-associated results for only autosomal sites [33]. For model details, see Materials and Methods.

**Figure 2.**
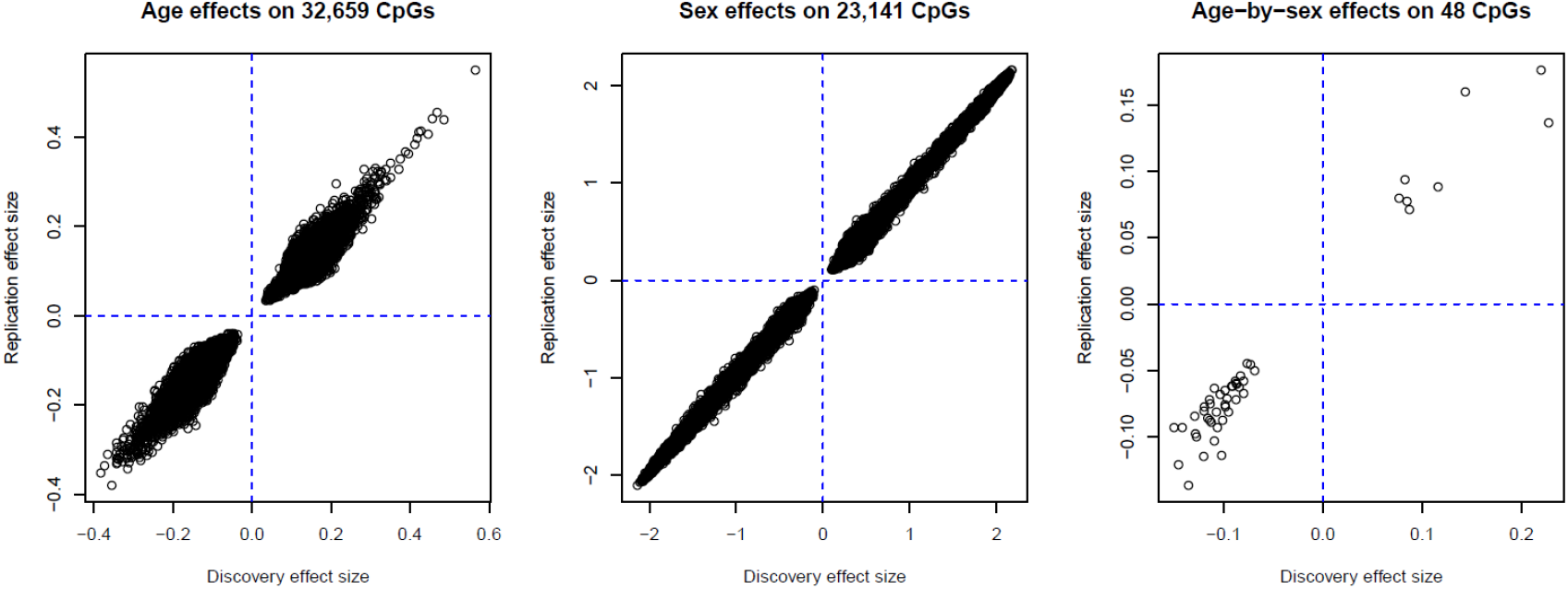
Age, sex, and age-by-sex effects in MCCS discovery and replication sets.

We further examined the replicated methylation markers using summary statistics results from two datasets from the Generation Scotland (GS) study [33]. We found that 94% age-associated (P<1.8×10^−6^), 86% sex-associated (P<5.1×10^−6^) and 91% age-by-sex-associated (P<0.0015) CpGs were replicated in GS data (Figure 1). The 30 replicated age-by-sex methylation markers (1 autosomal and 29 X-linked sites) are shown in Table 2.

**Table 2.** Thirty age-by-sex associations common to discovery (P<1.0×10^−7^) and replication (P<6.9×10^−4^) sets in MCCS and also replicated in GS data sets.

| CpG | CHR | MAPINFO | Strand | Gene | Gene feature | Discovery set in MCCS |  |  | Replication set in MCCS |  |  |
| --- | --- | --- | --- | --- | --- | --- | --- | --- | --- | --- | --- |
|  |  |  |  |  |  | Effect | SE | P value | Effect | SE | P value |
| cg12054453 | 17 | 57915717 | F | *TMEM49* | Body | 0.22 | 0.03 | 1.81E-12 | 0.18 | 0.03 | 5.56E-08 |
| cg06758848 | X | 153603268 | F | *FLNA* | TSS1500 | 0.14 | 0.02 | 9.77E-15 | 0.16 | 0.02 | 3.41E-16 |
| cg13775533 | X | 99663073 | F | *PCDH19* | 1stExon | -0.14 | 0.02 | 2.55E-14 | -0.09 | 0.02 | 8.69E-07 |
| cg09761247 | X | 148585951 | R | *IDS* | Body | 0.23 | 0.03 | 6.57E-13 | 0.14 | 0.03 | 2.96E-05 |
| cg08783090 | X | 151143125 | R | *GABRE* | 1stExon | -0.12 | 0.02 | 7.85E-13 | -0.08 | 0.02 | 3.27E-06 |
| cg25888700 | X | 21676344 | R | *KLHL34* | 5'UTR | -0.11 | 0.02 | 2.96E-12 | -0.08 | 0.02 | 3.68E-07 |
| cg01828474 | X | 21676593 | R | *KLHL34* | TSS200 | -0.11 | 0.02 | 8.17E-12 | -0.09 | 0.02 | 1.45E-07 |
| cg17036062 | X | 25034037 | F | *ARX* | 1stExon | -0.09 | 0.01 | 9.43E-12 | -0.06 | 0.01 | 1.20E-05 |
| cg25988710 | X | 72667379 | R | *CDX4* | 1stExon | -0.11 | 0.02 | 2.47E-11 | -0.07 | 0.02 | 3.18E-05 |
| cg06775759 | X | 21676994 | R | *KLHL34* | TSS1500 | -0.10 | 0.02 | 4.24E-11 | -0.09 | 0.02 | 2.92E-08 |
| cg03671371 | X | 36975540 | R | *NA* | *NA* | -0.11 | 0.02 | 4.72E-11 | -0.09 | 0.02 | 1.34E-06 |
| cg08814148 | X | 118407645 | F | *NA* | *NA* | -0.14 | 0.02 | 1.86E-10 | -0.14 | 0.02 | 1.57E-10 |
| cg16108684 | X | 125300035 | F | *DCAF12L2* | TSS200 | -0.12 | 0.02 | 3.66E-10 | -0.08 | 0.02 | 4.02E-05 |
| cg24823082 | X | 38660587 | F | *MID1IP1* | TSS200 | -0.15 | 0.02 | 4.31E-10 | -0.09 | 0.03 | 2.79E-04 |
| cg25156485 | X | 64887827 | R | *MSN* | Body | 0.08 | 0.01 | 5.02E-10 | 0.08 | 0.01 | 1.97E-10 |
| cg04424215 | X | 111325143 | R | *TRPC5* | 5'UTR | -0.13 | 0.02 | 6.53E-10 | -0.08 | 0.02 | 1.18E-04 |
| cg25528646 | X | 151143302 | R | *GABRE* | TSS200 | -0.09 | 0.01 | 8.18E-10 | -0.06 | 0.02 | 4.62E-05 |
| cg24931094 | X | 119737891 | R | *MCTS1* | 5'UTR | 0.09 | 0.01 | 1.31E-09 | 0.07 | 0.01 | 1.12E-06 |
| cg12537796 | X | 106515818 | R | *NA* | *NA* | -0.12 | 0.02 | 1.86E-09 | -0.09 | 0.02 | 8.22E-06 |
| cg25127732 | X | 21676692 | F | *KLHL34* | TSS1500 | -0.08 | 0.01 | 2.71E-09 | -0.05 | 0.01 | 3.56E-04 |
| cg03202526 | X | 139587311 | F | *SOX3* | TSS200 | -0.07 | 0.01 | 3.62E-09 | -0.05 | 0.01 | 1.52E-05 |
| cg11194545 | X | 105066793 | F | *NRK* | 5'UTR | -0.10 | 0.02 | 4.06E-09 | -0.08 | 0.02 | 9.14E-07 |
| cg06779802 | X | 107979401 | F | *IRS4* | 1stExon | -0.08 | 0.01 | 7.73E-09 | -0.07 | 0.01 | 1.30E-06 |
| cg22606540 | X | 100914483 | R | *ARMCX2* | 5'UTR | -0.10 | 0.02 | 1.23E-08 | -0.07 | 0.02 | 3.86E-05 |
| cg21729122 | X | 25034488 | F | *ARX* | TSS1500 | -0.10 | 0.02 | 1.34E-08 | -0.08 | 0.02 | 1.55E-05 |
| cg02295369 | X | 117959263 | R | *ZCCHC12* | Body | -0.09 | 0.02 | 3.41E-08 | -0.06 | 0.02 | 2.28E-04 |
| cg23208903 | X | 30327487 | F | *NR0B1* | 5'UTR | -0.09 | 0.02 | 3.82E-08 | -0.06 | 0.02 | 2.10E-04 |
| cg14586560 | X | 133118453 | R | *GPC3* | Body | -0.08 | 0.01 | 4.43E-08 | -0.06 | 0.01 | 8.19E-05 |
| cg21040569 | X | 137793665 | F | *FGF13* | Body | -0.08 | 0.02 | 5.24E-08 | -0.05 | 0.02 | 5.55E-04 |
| cg09407917 | X | 130192151 | R | *FLJ30058* | TSS200 | -0.09 | 0.02 | 6.52E-08 | -0.07 | 0.02 | 2.52E-05 |

### 2.3. Cancer risk

Of the 32,659 age-related methylation sites, two (cg25119261 in *HLA-DPB2* and cg05497216 in *ANKRD11*) were associated with risk of cancer overall (P<1.5×10^−6^ = 0.05/32659), and six (overlapping *AHSP, SBK1, RASGRP4* and *DPEP3* genes) were associated with risk of mature B-cell lymphoma (P<1.5×10^−6^) (Table 3). These CpGs were all replicated in GS data. Methylation M-values at these CpGs were both 1) all negatively associated with cancer risk (Table 3) and 2) all negatively associated with age (Supplementary Table 2). We found no evidence to support that methylation at any of the 48 CpGs showing age-by-sex interaction was associated with risk of cancer overall or specific type (P>0.001 = 0.05/48). A potential age-by-sex-related CpG, cg26738106 (chrX:3265038, TSS1500 of *MXRA5*), detected in the discovery set (beta=0.09 and P=3.0×10^−9^ in Model 2.2) was associated with colorectal cancer risk with OR=0.58 (95%CI: 0.44-0.77), P=0.0001 in Model 3.1, and OR=0.59 (95%CI: 0.44-0.78), P=0.0002 after adjustment for smoking, alcohol consumption and BMI (Model 3.2). However, this CpG was not replicated as showing age-by-sex-related methylation in the set of MCCS cases (beta=0.01 and P=0.36 in Model 2.2) and not included in the EPIC array used by McCartney et al. [33]’s GS study.

**Table 3.**
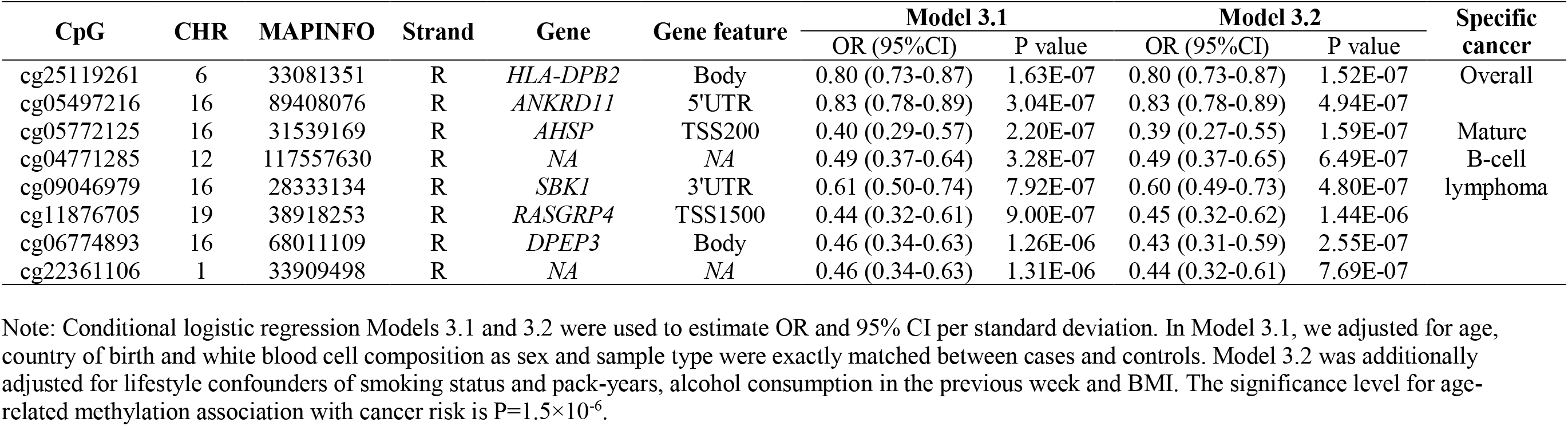
Significant CpGs for age associations with risk of cancer.

We further examined the trend of the association between epigenetic drift and risk of overall and specific cancers using the 32,659 age-associated CpGs. Figure 3 shows the correlations between the regression coefficients of the association between DNA methylation and age and the logarithm of ORs for the association between DNA methylation and cancer risk (the former was calculated in the discovery set). There was a strong positive correlation (beta=0.17, P<2.2×10^−16^) between associations of methylation with age and associations with overall cancer risk; the correlation was also strongly positive for kidney (beta=0.04, P=2.4×10^−8^), lung (beta=0.13, P<2.2×10^−16^) and urothelial (beta=0.33, P<2.2×10^−16^) cancers and mature B-cell lymphoma (beta=0.88, P<2.2×10^−16^); a negative correlation was observed for colorectal (beta=-0.09, P<2.2×10^−16^) and prostate (beta=-0.12, P<2.2×10^−16^) cancers.

**Figure 3.**
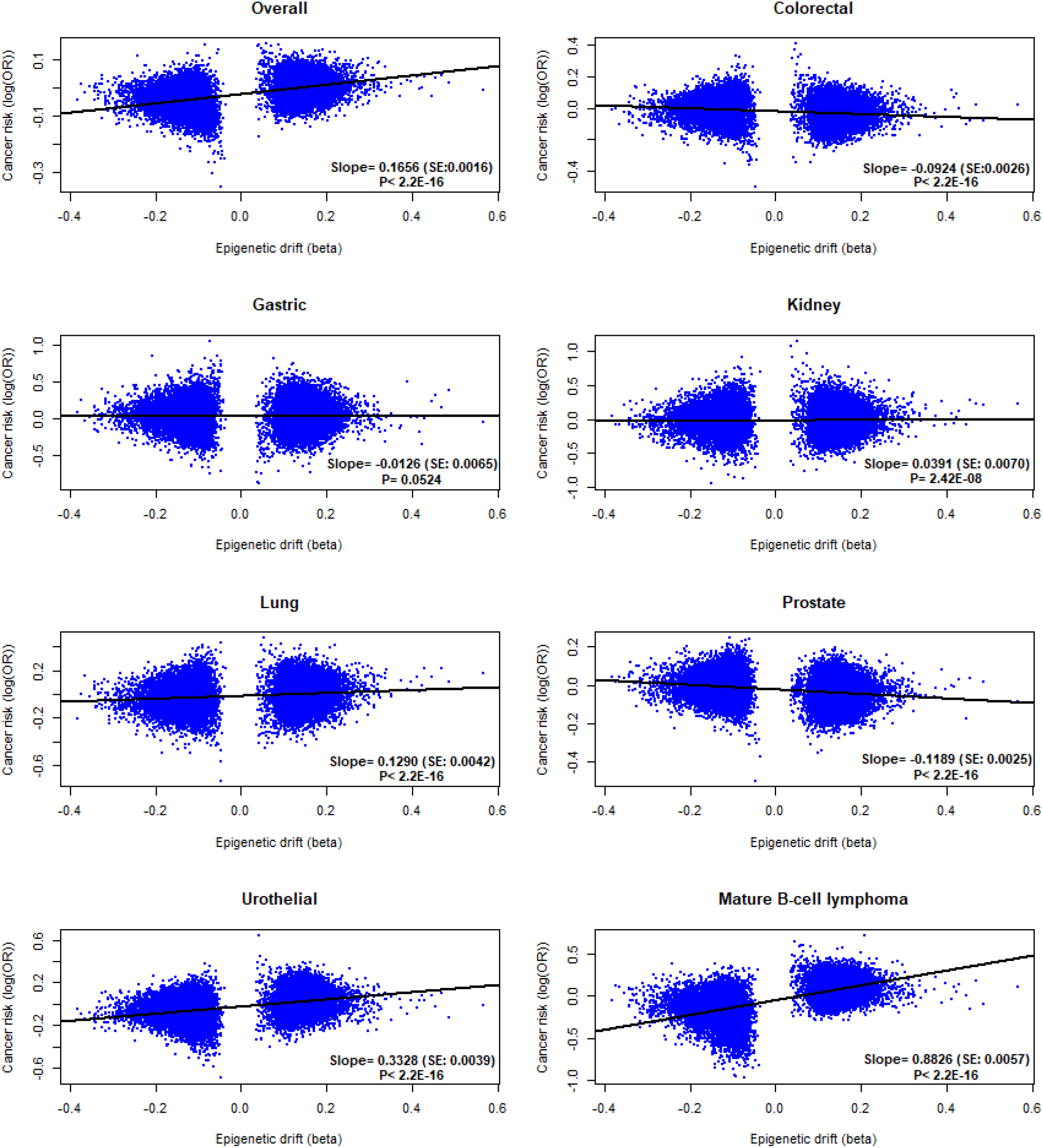
The trend of epigenetic drift with risk of different cancer types. Linear regression Y ∼ X was performed based on the 32,659 age-associated CpGs, where Y is log (OR) of DNA methylation association with risk of cancers and X is regression coefficient (beta) of DNA methylation association with age. Slope of the regression line, standard error (SE) and P value were obtained in the linear regression.

### 2.4. Cancer survival

Of the 32,659 age-related CpGs, 126 (all autosomal) were associated with survival of cancer overall (P<1.5×10^−6^) (Supplementary Table 5). The results for the 20 most significant associations are presented in Table 4. One age-by-sex-associated CpG cg12054453 (chr17:57915717) in the gene body of *TMEM49* was associated with survival of cancer overall (P<0.001) with HR=0.88 (95%CI: 0.83-0.93), P=2.3×10^−6^ in Model 4.1 and HR=0.91 (95%CI: 0.86-0.96), P=7.7×10^−4^ after adjustment for lifestyle-related confounders (Model 4.2).

**Table 4.** Top 20 significant CpGs for age associations with survival of cancer overall.

| CpG | CHR | MAPINFO | Strand | Gene | Gene feature | Model 4.1 |  | Model 4.2 |  |
| --- | --- | --- | --- | --- | --- | --- | --- | --- | --- |
|  |  |  |  |  |  | HR (95%CI) | P value | HR (95%CI) | P value |
| cg26427498 | 7 | 105987258 | R |  |  | 0.78 (0.74-0.82) | 8.35E-19 | 0.80 (0.76-0.85) | 1.21E-14 |
| cg26470501 | 19 | 45252955 | F | *BCL3* | Body | 0.80 (0.76-0.84) | 6.87E-18 | 0.84 (0.80-0.89) | 6.68E-11 |
| cg25143652 | 20 | 62168670 | R | *PTK6* | 1stExon | 0.76 (0.72-0.81) | 1.56E-16 | 0.81 (0.76-0.87) | 3.20E-10 |
| cg01127300 | 22 | 38614796 | F |  |  | 0.82 (0.78-0.86) | 5.19E-16 | 0.87 (0.83-0.91) | 1.07E-08 |
| cg08857221 | 1 | 37941361 | R | *ZC3H12A* | Body | 0.76 (0.71-0.81) | 1.34E-15 | 0.78 (0.73-0.84) | 4.82E-11 |
| cg19572487 | 17 | 38476024 | R | *RARA* | 5'UTR | 0.81 (0.77-0.86) | 4.51E-14 | 0.87 (0.82-0.92) | 4.85E-07 |
| cg02773019 | 3 | 135684688 | F | *PPP2R3A* | 5'UTR | 1.21 (1.15-1.27) | 6.22E-14 | 1.20 (1.14-1.26) | 7.34E-13 |
| cg15114651 | 19 | 47289410 | F | *SLC1A5* | TSS1500 | 0.77 (0.72-0.83) | 7.16E-14 | 0.83 (0.78-0.89) | 2.30E-07 |
| cg07069636 | 16 | 30671749 | F |  |  | 0.79 (0.74-0.84) | 1.04E-13 | 0.85 (0.79-0.90) | 2.95E-07 |
| cg15962267 | 5 | 138612986 | F | *SNHG4* | Body | 0.77 (0.72-0.83) | 1.54E-13 | 0.81 (0.75-0.87) | 3.70E-08 |
| cg02584867 | 19 | 1861099 | R | *KLF16* | Body | 0.83 (0.79-0.87) | 1.56E-13 | 0.85 (0.81-0.89) | 5.84E-11 |
| cg09287933 | 6 | 33384473 | R | *CUTA* | Body | 0.75 (0.70-0.81) | 1.56E-13 | 0.77 (0.72-0.84) | 8.99E-11 |
| cg03519879 | 14 | 74227499 | R | *C14orf43* | 5'UTR | 0.77 (0.72-0.83) | 2.46E-13 | 0.82 (0.77-0.88) | 3.72E-08 |
| cg12170787 | 19 | 1130965 | R | *SBNO2* | Body | 0.76 (0.71-0.82) | 5.32E-13 | 0.80 (0.74-0.86) | 5.26E-09 |
| cg07148697 | 6 | 31323253 | R | *HLA-B* | Body | 0.76 (0.71-0.82) | 5.67E-13 | 0.77 (0.72-0.84) | 3.32E-11 |
| cg26283141 | 6 | 33240471 | F | *VPS52* | TSS1500 | 0.82 (0.77-0.86) | 9.04E-13 | 0.84 (0.79-0.89) | 9.42E-10 |
| cg25264101 | 19 | 14064374 | F | *PODNL1* | TSS200 | 0.84 (0.80-0.88) | 1.29E-12 | 0.86 (0.82-0.90) | 3.27E-10 |
| cg05352838 | 6 | 33384391 | R | *CUTA* | 3'UTR | 0.73 (0.67-0.80) | 2.44E-12 | 0.76 (0.69-0.83) | 1.48E-09 |
| cg19513004 | 6 | 34206683 | F | *HMGA1* | 5'UTR | 0.80 (0.75-0.85) | 2.70E-12 | 0.83 (0.78-0.88) | 2.68E-09 |
| cg02003183 | 14 | 103415882 | F | *CDC42BPB* | Body | 1.21 (1.14-1.27) | 3.43E-12 | 1.17 (1.11-1.23) | 8.61E-09 |

Note: Cox regression Models 4.1 and 4.2 were used to estimate OR and 95% CI per standard deviation. In Model 4.1, we adjusted for age, sex, country of birth, sample type and white blood cell composition. Model 4.2 was additionally adjusted for smoking status and pack-years, alcohol consumption and BMI. The significance level for age-related methylation association with cancer risk is $P=1.5 \times 10^{-6}$ .

We also examined associations of age-related blood DNA methylation with survival for each individual cancer type and detected associations for gastric cancer (30 autosomal sites), lung cancer (282 autosomal and 1 X-linked sites) and mature B-cell lymphoma (1 autosomal site) (Supplementary Table 6). There was one age-by-sex-related CpG (cg21729122) in the TSS1500 region of *ARX* (X chromosome) that was associated with survival of colorectal cancer with HR=1.59 (95%CI: 1.28-1.97), P=2.3×10^−5^ in Model 4.1 and HR=1.52 (95%CI: 1.22-1.89), P=1.8×10^−4^ in Model 4.2. There was no evidence to support an association of age-by-sex-associated methylation sites with survival of other cancer types.

## 3. Discussion

Aging, as the main risk factor for most cancer types, captures the effects of cumulative exposure to exogenous and endogenous risk factors in one’s life time, thus the epigenetic drift reflects molecular alterations caused by both genetic and environmental risk factors [21]. Several studies [33-35] have investigated whether DNA methylation changes with age were different in males and females, using heterogeneous methods, data sources, and sample sizes. To our knowledge, our study using MCCS data is the largest single study to investigate cancer risk and survival with systematic, genome-wide assessment and replication of age and age-by-sex methylation signals. We found 32,659, 23,141 and 48 CpG sites at which methylation was strongly and consistently associated with age, sex and age-by-sex, respectively. These associations were all replicated internally, and their replication rate in the external data of GS [33] was as high as 94%, 86% and 91%, respectively. Our results for age-by-sex interaction (4 autosomal and 44 X-linked CpGs) corroborate the findings by McCartney et al. [33] that differences in age-associated DNA methylation between sexes are not frequent except for the X chromosome. Another recent study has indicated that DNA methylation patterns in the X-chromosome of whole blood leukocytes showed age- and sex-dependent with respect to X-inactivation [35].

Our results suggest that DNA methylation at some individual age-related CpGs is negatively associated with risk of cancer overall and mature B-cell lymphoma (Table 3). In the current study, DNA was extracted from pre-diagnostic peripheral blood, which may explain why comparatively more associations were observed for B-cell lymphoma compared with other types of cancer. Among these CpGs (Table 3), DNA methylation at cg25119261 (*HLA-DPB2*) was reported to be differentially methylated between tumor and matched adjacent normal tissues in the context of oral squamous cell [36] and hepatocellular cancer [37]. These conclusions may corroborate our finding of overall cancer association at this site, although we used DNA methylation from pre-diagnostic blood. Although the CpG cg26738106 (*MXRA5*) was negatively associated with risk of colorectal cancer in our data, the age-by-sex interaction observed in the set of controls was not replicated in the set of cases and therefore requires further investigation. It is however interesting to note that *MXRA5* was shown to be aberrantly expressed in colorectal tumor tissue [38]. A study using exome sequencing has also identified *MXRA5* as a cancer gene frequently mutated in non-small cell lung carcinoma [39], and the CpG cg26738106 in *MXRA5* was also nominally significantly associated (P<0.05) with survival from lung cancer in our data: HR=1.59 (95%CI: 1.17-2.15), P=0.003 in Model 4.1 and HR=1.58 (95%CI: 1.16-2.14), P=0.004 in Model 4.2.

One age-by-sex-associated CpG cg12054453 in *TMEM49* (also known as VMP1) was negatively associated with overall cancer survival. *TMEM49* has been widely reported as a cancer-relevant cell cycle modulator and its expression regulates the invasion and metastatic potential of cancer cells [40-42]. *ARX* is a homeobox-containing gene which is expressed primarily in the central and/or peripheral nervous system. Our study suggests that blood DNA methylation at cg21729122, located in the TSS1500 region of *ARX* may be associated with sex-specific survival from colorectal cancer. Although these findings should be interpreted with caution given methylation was measured in different tissues, they altogether suggest potential for blood DNA methylation in this region to provide a sex-specific mode of early detection and prediction of cancer.

Our results also showed a positive trend in the association between epigenetic drift and risk of cancer overall (slope=0.17) and cancers of the kidney (slope=0.04), lung (slope=0.13), and urothelium (slope=0.33) and mature B-cell lymphoma (slope=0.88), as well as a negative trend for gastric (slope= −0.01), prostate (slope= −0.12) and colorectal (slope= −0.09) cancers (Figure 3). This indicates that while our study may have been underpowered to detect associations at individual CpG sites, there is a strong link between aging of the blood methylation and risk for cancer. In two recent studies [28, 29], we investigated, using the same samples, the association between cancer risk and several synthetic measures of epigenetic aging. Most associations were overall similar but stronger for the second-generation measures such as *PhenoAge*, a composite biomarker of mortality, and *GrimAge*, a predictor of lifespan generated using several DNA methylation surrogates for plasma proteins and smoking history [24]. For *GrimAge*, we found that associations were in the same direction as the epigenetic drift (current results) with risk of most cancer types: overall (OR=1.11), kidney (OR=1.28), lung (OR=2.03), urothelial (OR=1.22), gastric (OR=0.95), prostate (OR=0.84) cancers and mature B-cell lymphoma (OR=1.03) [29]. We however observed that for colorectal cancer, the association of epigenetic drift with risk was in opposite direction as with *GrimAge* (OR=1.12), *PhenoAge* (OR=1.18) [29] and ‘first-generation’ epigenetic clocks [28]. This finding requires further investigation but could reflect biological differences between epigenetic drift and epigenetic clocks [21]. For prostate cancer, the strong negative tendency might reflect the advantaged background of men widely diagnosed with the disease via increased surveillance and testing in Australia.

There are several limitations in this study. First, there might have been a small proportion of closely related individuals between discovery and replication samples, which may result in some inflation of replicated signals. The fact that controls and cases were sampled from the same population and were all processed at the same time and using the same normalisation pipeline may also result in somewhat overconfident replication, compared with completely independent samples. Assessing associations using the set of cases may also result in collider bias, but it would likely be small in our setting given the matching of cases and controls [43, 44]. Secondly, the external GS data used the EPIC methylation array, which does not capture all the CpGs in our study using HM450; nevertheless, the vast majority of signals were replicated, so we anticipate that methylation sites that did not overlap would show similar replication rates. Despite these potential issues, our replication strategy was overall very stringent, as it used twice the Bonferroni correction for multiple testing, and showed very high consistency of the CpGs associated with age, sex, and age-by-sex. Thirdly, the sample sizes for investigating risk of and survival from specific cancers might not have been sufficiently large (only hundreds of each), to estimate associations with very good precision. It is therefore possible that associations were not detected by our study, but these would presumably be relatively weak and could be established via pooling our results with other methylation studies with similar design. Although little evidence of associations was found in the analyses of cancer risk and survival overall (all types), these analyses grouped together distinct cancer types, so might not be biologically relevant. Finally, we could not include a replication phase for the analyses of cancer risk and survival, so our findings of potential cancer biomarkers should be replicated in other studies, including mechanistic studies.

## 4. Materials and Methods

### 4.2. Study sample

The Melbourne Collaborative Cohort Study (MCCS) is an Australian prospective cohort study of 41,513 people of white European descent recruited between 1990 and 1994 in Melbourne metropolitan area. DNA was extracted from pre-diagnostic peripheral blood taken at recruitment (1990-1994) or at a subsequent follow-up visit (2003-2007) in cancer-free participants. More details about the cohort profile, blood collection, DNA extraction and cancer ascertainment can be found elsewhere [45]. Here we used a total of 3,215 case-control pairs from seven specific studies of colorectal (*N*=835), gastric (*N*=170), kidney (*N*=143), lung (*N*=332), prostate (*N*=869) and urothelial (*N*=428) cancers, and mature B-cell lymphoma (*N*=438) nested within the MCCS. Cases and controls were matched on age at blood draw, sex, country of birth (Australia/New-Zealand, Greece, Italy, or United Kingdom/other) and sample type (peripheral blood mononuclear cells, dried blood spots or buffy coats) using incidence density sampling. To minimise batch effects, samples from each matched case-control pair were plated to adjacent wells on the same BeadChip microarray, with plate, chip and position assigned randomly. Case-control pairs with missing values for the confounders (smoking status [current/ former/ never] and pack-years (log-transformed), alcohol consumption in the previous week [in grams/day] and body mass index [in kg/m^2^]) were excluded. All participants provided informed consent in accordance with the Declaration of Helsinki and the study was approved by Cancer Council Victoria’s Human Research Ethics Committee.

### 4.3. Quality control of DNA methylation data

DNA methylation in the MCCS was measured using the Illumina HumanMethylation450 (HM450) assay. Quality control (QC) details for processing methylation beta values have been reported previously [28]. Briefly, we removed CpGs with missing rate > 20% based on the sample and CpGs on Y-chromosome. M-values, calculated as 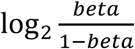, were then used for the analyses since these are thought to be more statistically valid for detection of differential methylation [46].

### 4.3. Statistical analyses

#### 4.3.1. Discovery and replication sets

To identify age-, sex- and age-by-sex-related methylation markers (Aims 1 and 2), we used all control subjects from the MCCS sample as a discovery set. After excluding duplicated controls across the seven studies (an individual may be assigned as a control in several different studies), 3,008 participants were available for the discovery phase analysis. After QC of methylation data on the discovery sample, there remained 484,828 available CpGs. All study cases were then used as the replication set. After excluding duplicates across the seven studies (an individual may be diagnosed with more than one specific cancer type and thus was selected as a case in several studies; in such instances, we included the first diagnosis only) and participants with samples that overlapped with the discovery set (an individual with a specific cancer may be assigned as a control in a different study), 2,754 participants were available for the replication phase analysis.

McCartney et al. [33] recently studied age-, sex- and age-by-sex-associated CpGs using DNA methylation data of Illumina EPIC array and samples (2,586 individuals for discovery and 4,450 individuals for replication) from Generation Scotland (GS) - Scottish Family Health Study. We used their summary statistics [47] as an external replication set.

#### 4.3.2. Age, sex, and age-by-sex associations

To assess associations of age and sex with DNA methylation (Aim 1), we conducted epigenome-wide association studies (EWAS). We fitted linear mixed regression models of methylation M-values at individual CpGs on age, sex, country of birth and sample type as fixed effect variables, and study of specific cancer type, assay plate and slide as random effect variables. A first model (Model 1.1) was adjusted for white blood cell composition (percentage of CD4 + T cells, CD8 + T cells, B cells, NK cells, monocytes and granulocytes, estimated using the Houseman algorithm [48]), and a second model (Model 1.2) was additionally adjusted for lifestyle factors (smoking status and pack-years, alcohol consumption in the previous week and BMI [49-51]). Age-associated and sex-associated changes in DNA methylation were then assessed by examining regression coefficients for the variables age and sex, respectively.

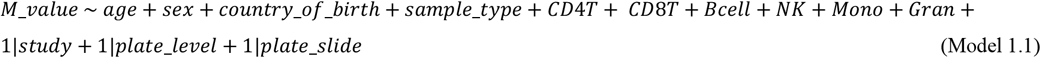

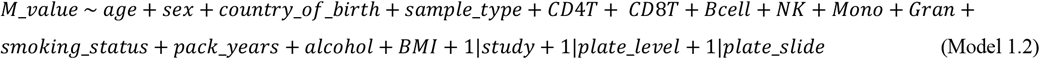

To investigate sex-specific ageing-associated changes in DNA methylation (Aim 2), we added an interaction term age × sex to the same models (Model 2.1 and Model 2.2).

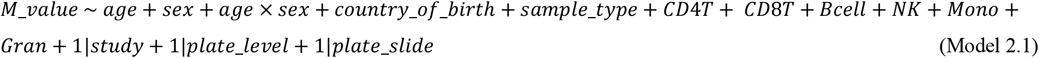

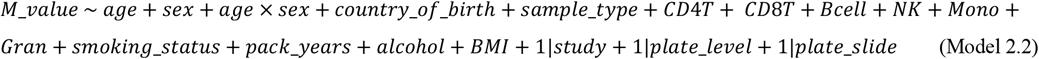

In the discovery phase, we used the Bonferroni correction for multiple testing to declare statistical significance (P<0.05/484828=1.0×10^−7^) for all Model 1s and 2s. The same models were then carried out using the replication set. Replicated findings were further examined using the GS summary statistics [33,45] (considering only CpGs common to the HM450 and EPIC assays). The replication criteria included multiple comparisons (P-value threshold for replication was based on Bonferroni correction) and same effect directions. A flowchart detailing the study design and analysis pipeline was presented in Figure 1.

#### 4.3.3. Cancer risk and survival associations

We assessed associations between DNA methylation at individual age- and age-by-sex-associated CpGs and risk of overall cancer (using the 2,754 matched case-control pairs with no duplicated participant involved) and cancer at seven specific sites, using conditional logistic regression models (Models 3.1 and 3.2) to estimate odds ratio (OR) and 95% confidence intervals (CI), expressed per standard deviation (SD). In Model 3.1, we adjusted for age, country of birth and white blood cell composition. Sex and sample type were exactly matched between cases and controls so were not adjusted for. Model 3.2 was additionally adjusted for smoking status (current / former / never) and pack-years, alcohol consumption in the previous week (grams/day, continuous) and BMI (continuous). The methylation sites at which associations were significant after Bonferroni correction for multiple comparisons in both Models 3.1 and 3.2 were considered to be associated with risk of cancer.

We used Cox models (Models 4.1 and 4.2) to estimate hazard ratios (HR) per SD for the associations between M-values at individual age- and age-by-sex-associated CpG sites and risk of death (all causes) following cancer diagnosis. The survival analysis was thus restricted to cancer cases. A total of *N*=1,931 deaths were included in the analysis. Time since diagnosis was used as the timescale, and person-years of follow-up were calculated from the diagnosis date until the date of death, and censored at the date of departure from Australia or end of follow-up. Where a participant was diagnosed with several cancers, we considered only the first diagnosis to count follow-up time. Number of deaths by cancer type was as follows: colorectal cancer: *N*=526, gastric cancer: *N*=144, kidney cancer: *N*=80, lung cancer: *N*=311, prostate cancer: *N*=435, urothelial cancer: *N*=234, and mature B-cell lymphoma: *N*=286. In Model 4.1, we adjusted for age, sex, country of birth, sample type and white blood cell composition. Model 4.2 was additionally adjusted for smoking status and pack-years, alcohol consumption and BMI. The methylation sites at which associations were Bonferroni-significant in both Models 4.1 and 4.2 were considered to be associated with cancer survival. All statistical analyses were performed using R version 3.6.0.

## 5. Conclusions

Our study is the first large-scale study to examine potential genes at which blood DNA methylation is age-related (including modification by sex) and associated with cancer development and progression. Our results may be useful for developing strategies of early diagnosis of cancer and prediction of cancer prognosis.

## Data Availability

Data will be made available upon reasonable request to the corresponding author.

## Supplementary Materials

The following are available online at the website of the journal.

**Figure S1**: Associations of methylation M-values with age-by-sex for the 48 CpGs.

**Table S1**. Characteristics of the participants from seven specific studies in the MCCS sample.

**Table S2**: 32,659 replicated age-associated CpGs using the MCCS sample.

**Table S3**: 23,141 replicated sex-associated CpGs using the MCCS sample.

**Table S4**: 48 replicated age-by-sex-associated CpGs using the MCCS sample.

**Table S5**: 126 (all autosomal) age-related CpGs were associated with survival of cancer overall.

**Table S6**: Significant age-related CpGs with survival of gastric cancer (30 autosomal sites), lung cancer (282 autosomal and 1 X-linked) and mature B-cell lymphoma (1 autosomal site).

## Author Contributions

Conceptualization, C.Y. and P.A.D.; methodology, C.Y. and P.A.D.; validation, C.Y.; formal analysis, C.Y. and P.A.D.; investigation, C.Y. and P.A.D.; resources, G.G.G. and M.C.S.; laboratory measurements, E.M.W. and J.E.J; data curation, E.E.M., J.E.J, C.Y. and P.A.D.; Writing-Original draft preparation, C.Y. and P.A.D.; Writing-Review and editing, all authors; visualization, C.Y.; supervision, P.A.D.; project administration, M.C.S. and P.A.D.; funding acquisition, E.E.M., J.E.J., A.M.H., E.M., D.S., D.D.B, G.S., D.R.E., G.G.G, M.C.S. and P.A.D. All authors have read and agreed to the published version of the manuscript.

## Institutional Review Board Statement

The study was approved by Cancer Council Victoria’s Human Research Ethics Committee, Melbourne, VIC, Australia.

## Informed Consent Statement

All participants provided informed consent in accordance with the Declaration of Helsinki.

## References

1. US Cancer Statistics Working Group, 2013. United States cancer statistics: 1999-2010 incidence and mortality web-based report. Atlanta: US Department of Health and Human Services, Centers for Disease Control and Prevention and National Cancer Institute, 201.

2. Campisi, J., 2013. Aging, cellular senescence, and cancer. Annual review of physiology, 75, 685–705.

3. White, M.C., Holman, D.M., Boehm, J.E., Peipins, L.A., Grossman, M. and Henley, S.J., 2014. Age and cancer risk: a potentially modifiable relationship. American journal of preventive medicine, 46(3), S7–S15.

4. Aunan, J.R., Cho, W.C. and Søreide, K., 2017. The biology of aging and cancer: a brief overview of shared and divergent molecular hallmarks. Aging and disease, 8(5), 628.

5. Cook, M.B., Dawsey, S.M., Freedman, N.D., Inskip, P.D., Wichner, S.M., Quraishi, S.M., Devesa, S.S. and McGlynn, K.A., 2009. Sex disparities in cancer incidence by period and age. Cancer Epidemiology and Prevention Biomarkers, 18(4), 1174–1182.

6. Cook, M.B., McGlynn, K.A., Devesa, S.S., Freedman, N.D. and Anderson, W.F., 2011. Sex disparities in cancer mortality and survival. Cancer Epidemiology and Prevention Biomarkers, 20(8), 1629–1637.

7. Edgren, G., Liang, L., Adami, H.O. and Chang, E.T., 2012. Enigmatic sex disparities in cancer incidence. European journal of epidemiology, 27(3), 187–196.

8. Zhu, Y., Shao, X., Wang, X., Liu, L. and Liang, H., 2019. Sex disparities in cancer. Cancer letters, 466, 35–38.

9. Lopes-Ramos, C.M., Quackenbush, J. and DeMeo, D.L., 2020. Genome-Wide Sex and Gender Differences in Cancer. Frontiers in Oncology, 10, 2486.

10. Rubin, J.B., Lagas, J.S., Broestl, L., Sponagel, J., Rockwell, N., Rhee, G., Rosen, S.F., Chen, S., Klein, R.S., Imoukhuede, P. and Luo, J., 2020. Sex differences in cancer mechanisms. Biology of sex Differences, 11, 1–29.

11. Majek, O., Gondos, A., Jansen, L., Emrich, K., Holleczek, B., Katalinic, A., Nennecke, A., Eberle, A., Brenner, H. and GEKID Cancer Survival Working Group, 2013. Sex differences in colorectal cancer survival: population-based analysis of 164,996 colorectal cancer patients in Germany. PloS one, 8(7), e68077.

12. Song, M., Kang, D., Yang, J.J., Choi, J.Y., Sung, H., Lee, Y., Yoon, H.S., Choi, Y., Kong, S.H., Lee, H.J. and Yang, H.K., 2015. Age and sex interactions in gastric cancer incidence and mortality trends in Korea. Gastric Cancer, 18(3), 580–589.

13. Qu, Y., Chen, H., Gu, W., Gu, C., Zhang, H., Xu, J., Zhu, Y. and Ye, D., 2015. Age-dependent association between sex and renal cell carcinoma mortality: a population-based analysis. Scientific reports, 5, 9160.

14. Zeng, C., Wen, W., Morgans, A.K., Pao, W., Shu, X.O. and Zheng, W., 2015. Disparities by race, age, and sex in the improvement of survival for major cancers: results from the National Cancer Institute Surveillance, Epidemiology, and End Results (SEER) Program in the United States, 1990 to 2010. JAMA oncology, 1(1), 88–96.

15. Mamtani, R., Wang, X.V., Gyawali, B., DiPaola, R.S., Epperson, C.N., Haas, N.B. and Dutcher, J.P., 2019. Association between age and sex and mortality after adjuvant therapy for renal cancer. Cancer, 125(10), 1637–1644.

16. Sedivy, J.M., Banumathy, G. and Adams, P.D., 2008. Aging by epigenetics - a consequence of chromatin damage? Experimental cell research, 314(9), 1909–1917.

17. Pal, S. and Tyler, J.K., 2016. Epigenetics and aging. Science advances, 2(7), e1600584.

18. Liu, J., Morgan, M., Hutchison, K. and Calhoun, V.D., 2010. A study of the influence of sex on genome wide methylation. PloS one, 5(4), e10028.

19. Khramtsova, E.A., Davis, L.K. and Stranger, B.E., 2019. The role of sex in the genomics of human complex traits. Nature Reviews Genetics, 20(3), 173–190.

20. Jones, M.J., Goodman, S.J. and Kobor, M.S., 2015. DNA methylation and healthy human aging. Aging cell, 14(6), 924–932.

21. Zheng, S.C., Widschwendter, M. and Teschendorff, A.E., 2016. Epigenetic drift, epigenetic clocks and cancer risk. Epigenomics, 8(5), 705–719.

22. Horvath, S., 2013. DNA methylation age of human tissues and cell types. Genome biology, 14(10), 3156.

23. Levine, M.E., Lu, A.T., Quach, A., Chen, B.H., Assimes, T.L., Bandinelli, S., Hou, L., Baccarelli, A.A., Stewart, J.D., Li, Y. and Whitsel, E.A., 2018. An epigenetic biomarker of aging for lifespan and healthspan. Aging (Albany NY), 10(4), 573.

24. Lu, A.T., Quach, A., Wilson, J.G., Reiner, A.P., Aviv, A., Raj, K., Hou, L., Baccarelli, A.A., Li, Y., Stewart, J.D. and Whitsel, E.A., 2019. DNA methylation GrimAge strongly predicts lifespan and healthspan. Aging (Albany NY), 11(2), 303.

25. Horvath, S. and Raj, K., 2018. DNA methylation-based biomarkers and the epigenetic clock theory of ageing. Nature Reviews Genetics, 19(6), 371.

26. Dugue, P. A., Bassett, J. K., Joo, J. E., Baglietto, L., Jung, C. H., Wong, E. M., Fiorito, G., Schmidt, D., Makalic, E., Li, S., Moreno-Betancur, M., Buchanan, D. D., Vineis, P., English, D. R., Hopper, J. L., Severi, G., Southey, M. C., Giles, G. G., Milne, R. L., Association of DNA methylation-based biological age with health risk factors and overall and cause-specific mortality. American journal of epidemiology 2018, 187, (3), 529–538.

27. Dugué, P.-A., Li, S., Hopper, J. L., Milne, R. L., Chapter 3 - DNA Methylation-Based Measures of Biological Aging. In Epigenetics in Human Disease (Second Edition), Tollefsbol, T. O., Ed. Academic Press: London, 2018; 6, 39–64.

28. Dugué, P.A., Bassett, J.K., Joo, J.E., Jung, C.H., Ming Wong, E., Moreno-Betancur, M., Schmidt, D., Makalic, E., Li, S., Severi, G. and Hodge, A.M., 2018. DNA methylation-based biological aging and cancer risk and survival: Pooled analysis of seven prospective studies. International journal of cancer, 142(8), 1611–1619.

29. Dugué, P.A., Bassett, J.K., Wong, E.M., Joo, J.E., Li, S., Yu, C., Schmidt, D.F., Makalic, E., Doo, N.W., Buchanan, D.D. and Hodge, A.M., 2021. Biological aging measures based on blood DNA methylation and risk of cancer: a prospective study. JNCI cancer spectrum, 5(1), pkaa109.

30. Chung, M., Ruan, M., Zhao, N., Koestler, D.C., De Vivo, I., Kelsey, K.T. and Michaud, D.S., 2021. DNA methylation ageing clocks and pancreatic cancer risk: Pooled analysis of three prospective nested case-control studies. Epigenetics, 1–11.

31. Ambatipudi, S., Horvath, S., Perrier, F., Cuenin, C., Hernandez-Vargas, H., Le Calvez-Kelm, F., Durand, G., Byrnes, G., Ferrari, P., Bouaoun, L. and Sklias, A., 2017. DNA methylome analysis identifies accelerated epigenetic ageing associated with postmenopausal breast cancer susceptibility. European journal of cancer, 75, 299–307.

32. Gào, X., Zhang, Y., Boakye, D., Li, X., Chang-Claude, J., Hoffmeister, M. and Brenner, H., 2020. Whole blood DNA methylation aging markers predict colorectal cancer survival: a prospective cohort study. Clinical epigenetics, 12(1), 1–13.

33. McCartney, D.L., Zhang, F., Hillary, R.F., Zhang, Q., Stevenson, A.J., Walker, R.M., Bermingham, M.L., Boutin, T., Morris, S.W., Campbell, A. and Murray, A.D., 2020. An epigenome-wide association study of sex-specific chronological ageing. Genome medicine, 12(1), 1–11.

34. Yusipov, I., Bacalini, M.G., Kalyakulina, A., Krivonosov, M., Pirazzini, C., Gensous, N., Ravaioli, F., Milazzo, M., Giuliani, C., Vedunova, M. and Fiorito, G., 2020. Age-related DNA methylation changes are sex-specific: a comprehensive assessment. Aging (Albany NY), 12(23), 24057.

35. Li, S., Lund, J.B., Christensen, K., Baumbach, J., Mengel-From, J., Kruse, T., Li, W., Mohammadnejad, A., Pattie, A., Marioni, R.E. and Deary, I.J., 2020. Exploratory analysis of age and sex dependent DNA methylation patterns on the X-chromosome in whole blood samples. Genome Medicine, 12, 1–13.

36. Khongsti, S., Lamare, F.A., Shunyu, N.B., Ghosh, S., Maitra, A. and Ghosh, S., 2018. Whole genome DNA methylation profiling of oral cancer in ethnic population of Meghalaya, North East India reveals novel genes. Genomics, 110(2), 112–123.

37. Sun, X.J., Wang, M.C., Zhang, F.H. and Kong, X., 2018. An integrated analysis of genome-wide DNA methylation and gene expression data in hepatocellular carcinoma. FEBS open bio, 8(7), 1093–1103.

38. Wang, G.H., Yao, L., Xu, H.W., Tang, W.T., Fu, J.H., Hu, X.F., Cui, L. and Xu, X.M., 2013. Identification of MXRA5 as a novel biomarker in colorectal cancer. Oncology letters, 5(2), 544–548.

39. Xiong, D., Li, G., Li, K., Xu, Q., Pan, Z., Ding, F., Vedell, P., Liu, P., Cui, P., Hua, X. and Jiang, H., 2012. Exome sequencing identifies MXRA5 as a novel cancer gene frequently mutated in non-small cell lung carcinoma from Chinese patients. Carcinogenesis, 33(9), 1797–1805.

40. Sauermann, M., Sahin, Ö., Sültmann, H., Hahne, F., Blaszkiewicz, S., Majety, M., Zatloukal, K., Füzesi, L., Poustka, A., Wiemann, S. and Arlt, D., 2008. Reduced expression of vacuole membrane protein 1 affects the invasion capacity of tumor cells. Oncogene, 27(9), 1320–1326.

41. Qian, Q., Zhou, H., Chen, Y., Shen, C., He, S., Zhao, H., Wang, L., Wan, D. and Gu, W., 2014. VMP1 related autophagy and apoptosis in colorectal cancer cells: VMP1 regulates cell death. Biochemical and biophysical research communications, 443(3), 1041–1047.

42. Zheng, L., Chen, L., Zhang, X., Zhan, J. and Chen, J., 2016. TMEM49-related apoptosis and metastasis in ovarian cancer and regulated cell death. Molecular and cellular biochemistry, 416(1-2), 1–9.

43. Greenland, S., 2003. Quantifying biases in causal models: classical confounding vs collider-stratification bias. Epidemiology, 300–306.

44. Pearce, N. and Richiardi, L., 2014. Commentary: three worlds collide: Berkson’s bias, selection bias and collider bias. International journal of epidemiology, 43(2), 521–524.

45. Milne, R.L., Fletcher, A.S., MacInnis, R.J., Hodge, A.M., Hopkins, A.H., Bassett, J.K., Bruinsma, F.J., Lynch, B.M., Dugué, P.A., Jayasekara, H. and Brinkman, M.T., 2017. Cohort profile: the Melbourne collaborative cohort study (health 2020). International journal of epidemiology, 46(6), pp.1757–1757i.

46. Du, P., Zhang, X., Huang, C.C., Jafari, N., Kibbe, W.A., Hou, L. and Lin, S.M., 2010. Comparison of Beta-value and M-value methods for quantifying methylation levels by microarray analysis. BMC bioinformatics, 11(1), p.587.

47. McCartney, Daniel; Zhang, Futao; Hillary, Robert; Zhang, Qian; Stevenson, Anna; Walker, Rosie; Bermingham, Mairead; Boutin, Thibaud; Morris, Stewart; Campbell, Archie; Murray, Alison; Whalley, Heather; Porteous, David; Hayward, Caroline; Evans, Kathy; Chandra, Tamir; Deary, Ian; McIntosh, Andrew; Yang, Jian; Visscher, Peter; McRae, Allan; Marioni, Riccardo. (2019). Generation Scotland Age x Sex Epigenome Wide Association Study Summary Statistics, [dataset]. University of Edinburgh. https://doi.org/10.7488/ds/2709.

48. Houseman, E.A., Accomando, W.P., Koestler, D.C., Christensen, B.C., Marsit, C.J., Nelson, H.H., Wiencke, J.K. and Kelsey, K.T., 2012. DNA methylation arrays as surrogate measures of cell mixture distribution. BMC bioinformatics, 13(1), p.86.

49. Dugue, P. A., Jung, C. H., Joo, J. E., Wang, X., Wong, E. M., Makalic, E., Schmidt, D. F., Baglietto, L., Severi, G., Southey, M. C., English, D. R., Giles, G. G., Milne, R. L., Smoking and blood DNA methylation: an epigenome-wide association study and assessment of reversibility. Epigenetics: official journal of the DNA Methylation Society 2020, 15, (4), 358–368.

50. Dugue, P. A., Wilson, R., Lehne, B., Jayasekara, H., Wang, X., Jung, C. H., Joo, J. E., Makalic, E., Schmidt, D. F., Baglietto, L., Severi, G., Gieger, C., Ladwig, K. H., Peters, A., Kooner, J. S., Southey, M. C., English, D. R., Waldenberger, M., Chambers, J. C., Giles, G. G., Milne, R. L., Alcohol consumption is associated with widespread changes in blood DNA methylation: Analysis of cross-sectional and longitudinal data. Addiction biology 2019, e12855.

51. Geurts, Y. M., Dugue, P. A., Joo, J. E., Makalic, E., Jung, C. H., Guan, W., Nguyen, S., Grove, M. L., Wong, E. M., Hodge, A. M., Bassett, J. K., FitzGerald, L. M., Tsimiklis, H., Baglietto, L., Severi, G., Schmidt, D. F., Buchanan, D. D., MacInnis, R. J., Hopper, J. L., Pankow, J. S., Demerath, E. W., Southey, M. C., Giles, G. G., English, D. R., Milne, R. L., Novel associations between blood DNA methylation and body mass index in middle-aged and older adults. Int J Obes (Lond) 2018, 42, (4), 887–896.

