## Supplementary figures and images for "Epigenetic drift association with cancer risk and survival, and modification by sex"

### Supplementary Figure 1

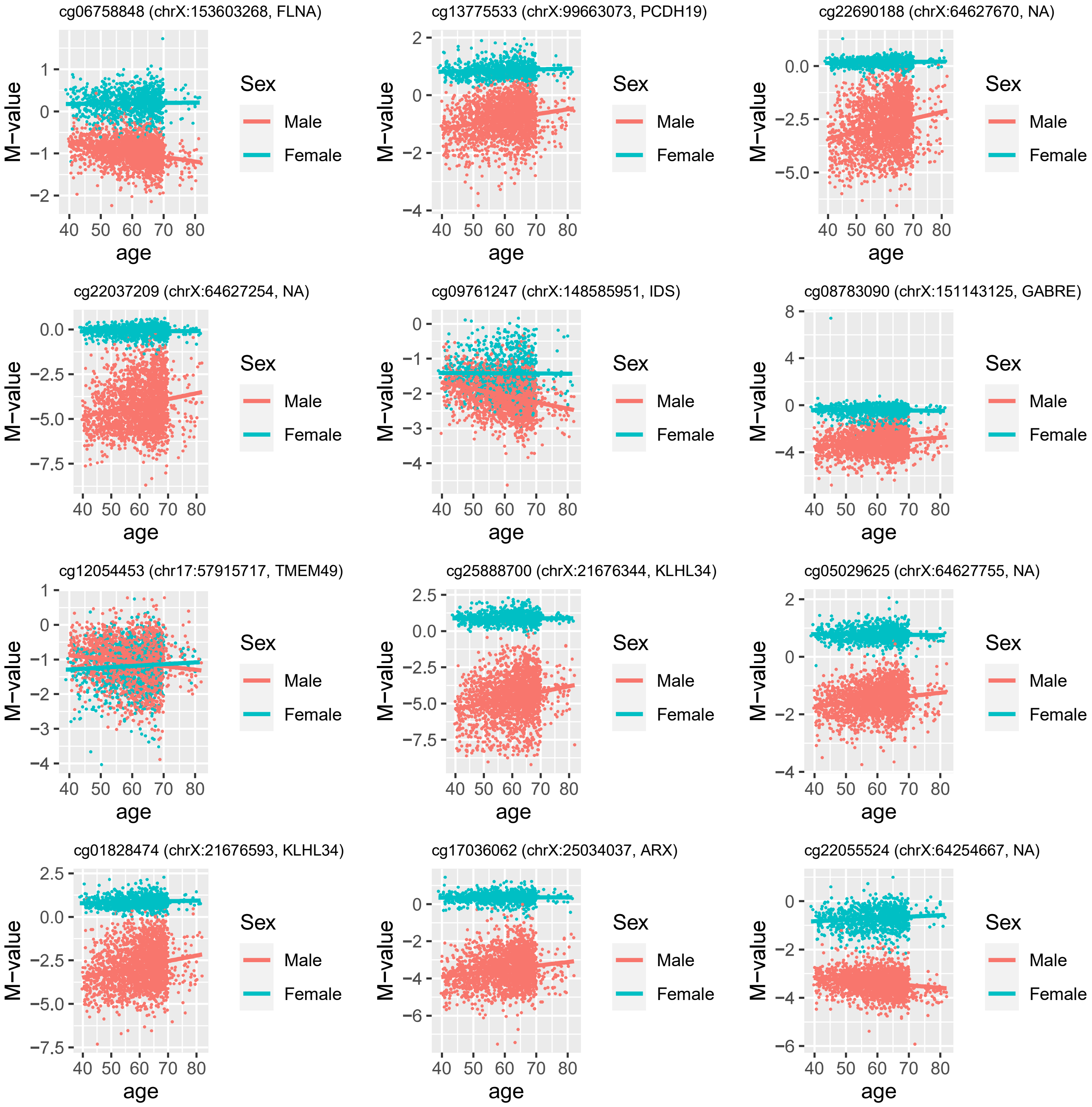

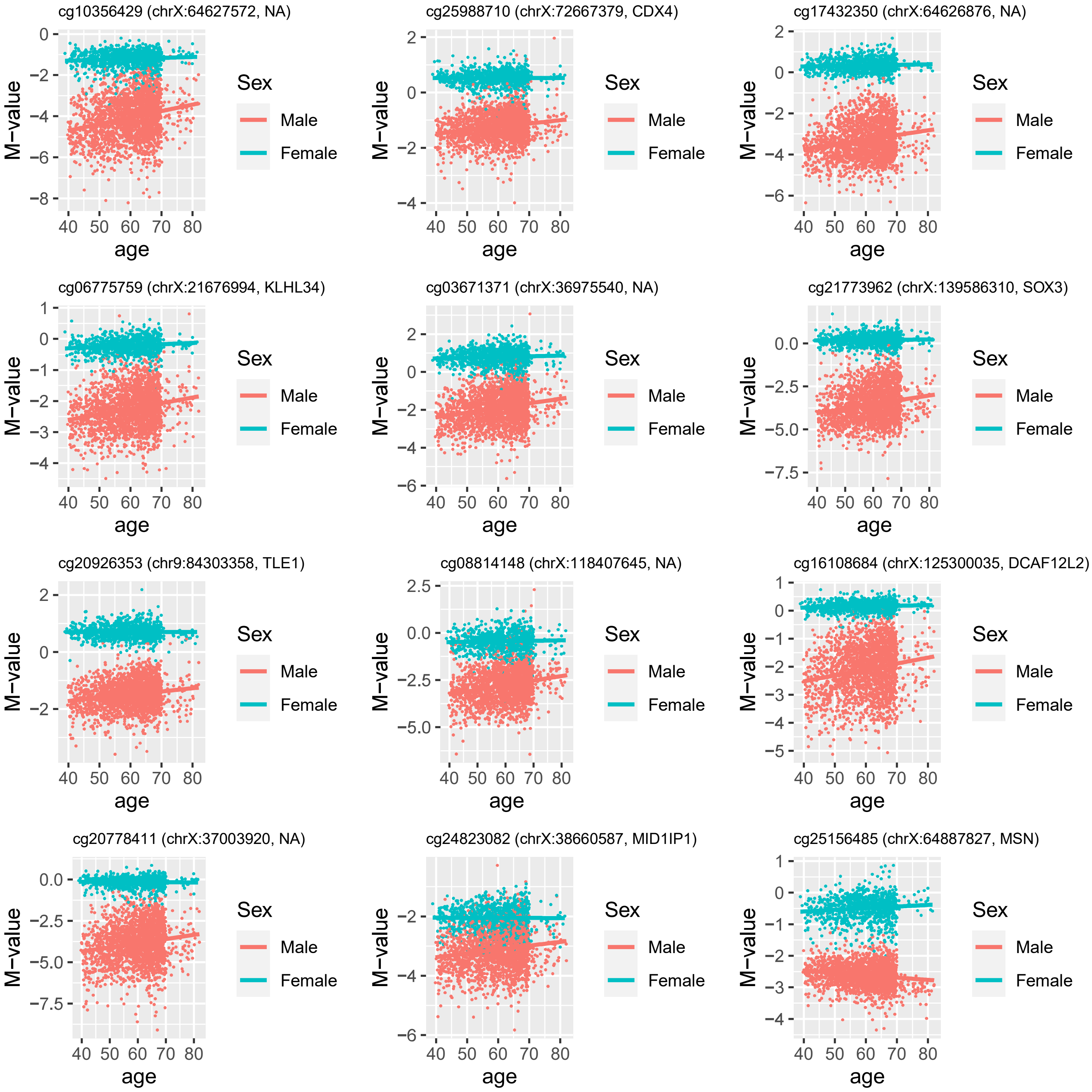

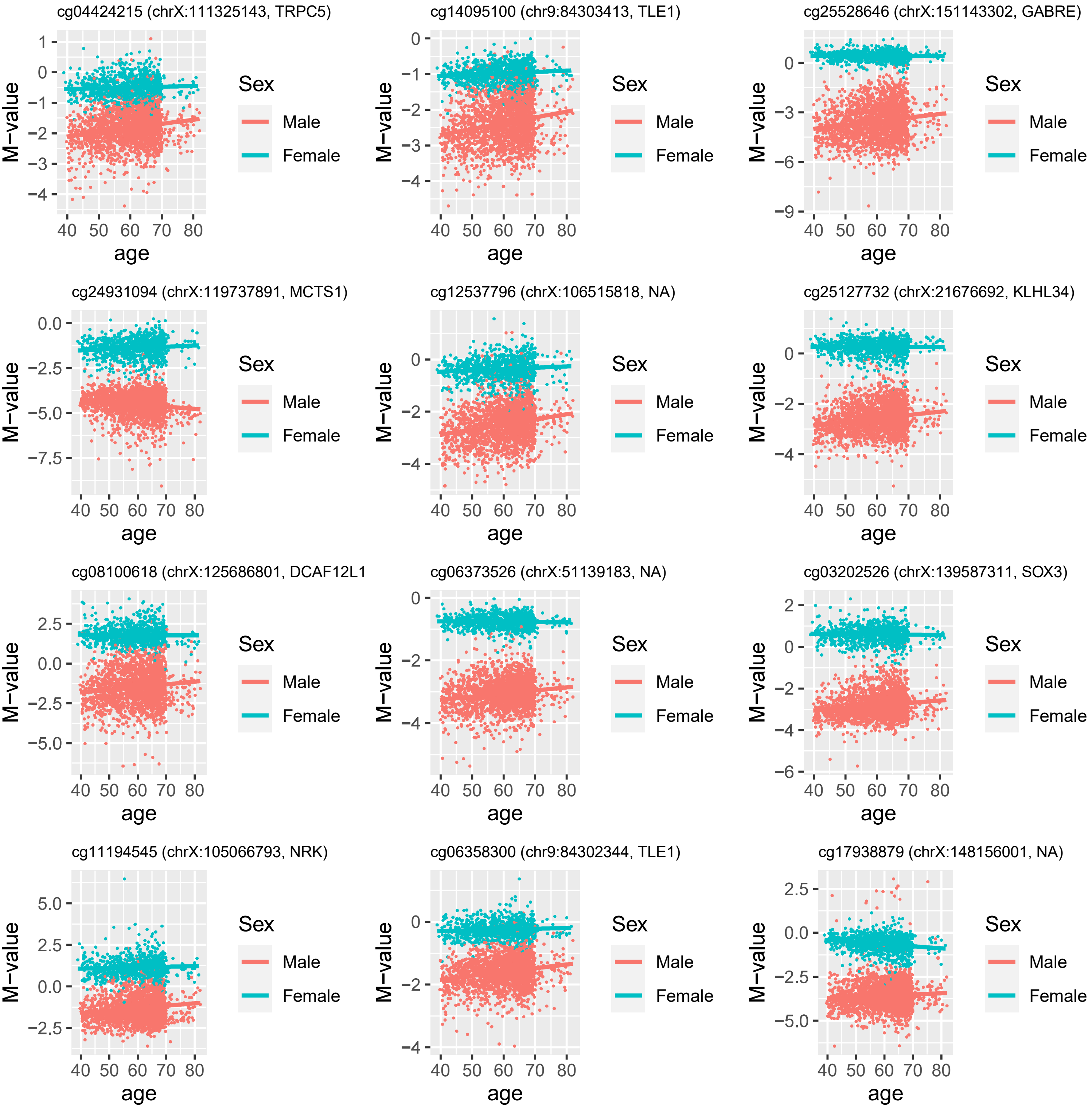

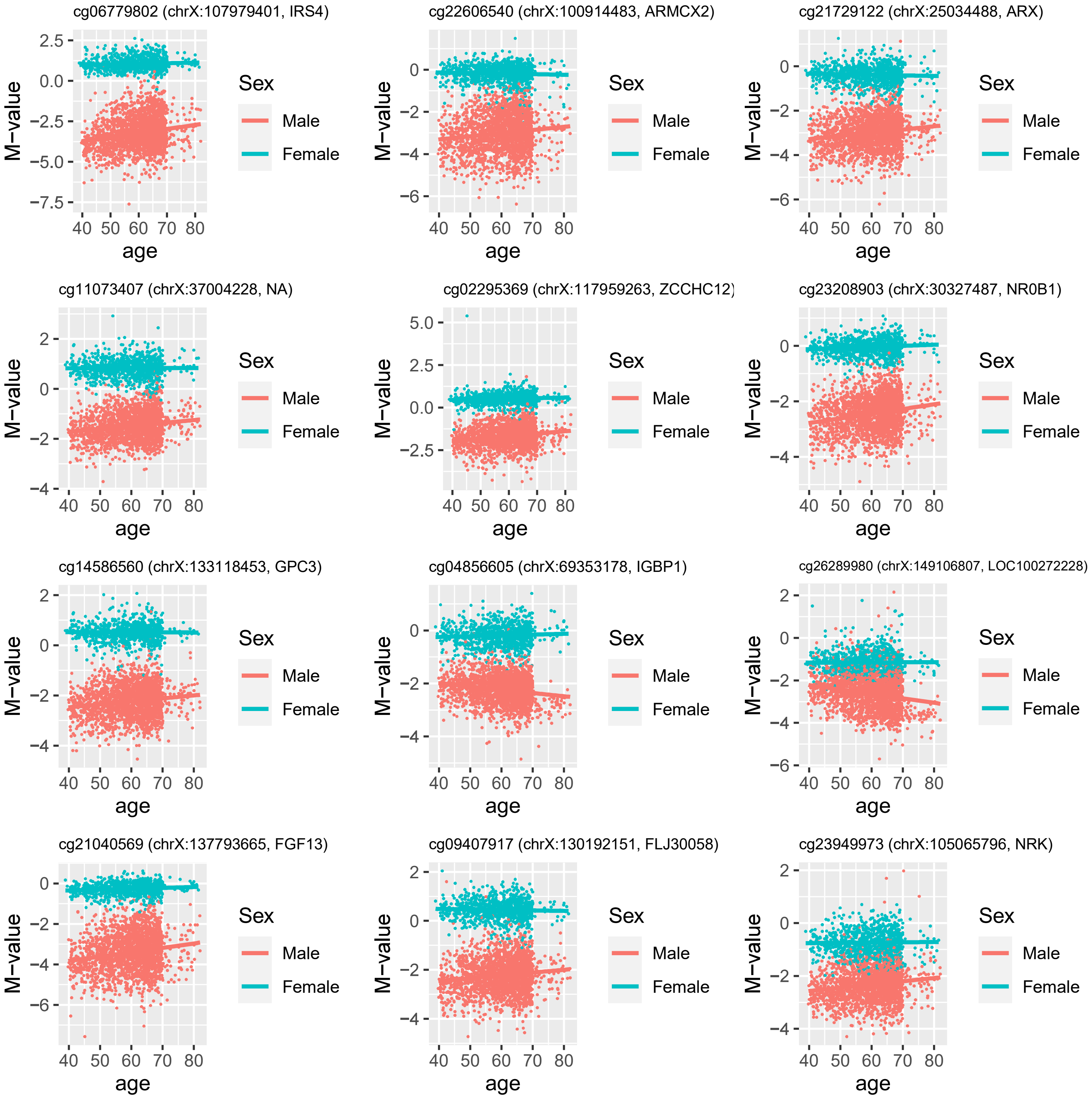
